# Pre-operative Spine Tumor Embolization: Clinical Outcomes and Effect of Embolization Completeness

**DOI:** 10.1101/2024.01.20.24301548

**Authors:** Nima Omid-Fard, Jean-Paul Salameh, Matthew DF McInnes, Charles G Fisher, Manraj KS Heran

## Abstract

**Background and Purpose:** To assess the association between the impact of the completeness of pre-operative spine tumour embolization and clinical outcomes including estimated blood loss (EBL), neurological status, and complications.

**Materials and Methods:** Retrospective chart review of all preoperative spine tumour embolization procedures performed over 11 years by a single operator (2007-2018) at Vancouver General Hospital, on 40 consecutive patients (mean age 58; 77.5% males) with 42 embolization procedures, of which surgery was done en bloc in 22 cases and intralesional in the remaining 20. A multivariable negative binomial regression model was fit to examine the association between EBL and surgery type, tumour characteristics, embolization completeness and operative duration.

**Results:** Among intralesional surgeries, complete versus incomplete embolization was associated with reduced blood loss (772 vs 1428 mL, P < 0.001). There was no statistically significant difference in neurological outcomes or complications between groups. Highly vascular tumours correlated with greater blood loss than their less vascular counterparts, but tumour location did not have a statistically significant effect.

**Conclusion:** This study provides early evidence in support of our hypothesis that complete as opposed to incomplete tumour embolization correlates with reduced blood loss in intralesional surgeries. Randomized control trials with larger samples are necessary to confirm this benefit and to ascertain other potential clinical benefits.

## Introduction

Preoperative embolization has become a routine procedure to treat spine neoplasms prior to resection, with proposed clinical benefits including reduced estimated blood loss (EBL), improved pain profile, and an ability to avoid deterioration in neurological status.^1-7^ Presumably, these effects may be achieved through faster, simpler surgeries that allow completion of planned procedures more often. However, the extent to which clinical benefits are attained depends on the tumour vascularity,^8, 9^ whether surgery is intralesional or en bloc,^5^ and the degree to which embolization is achieved.^10, 11^ The latter factor is of particular interest to the interventionist as it is technique and operator dependent. Nevertheless, the true impact of preoperative embolization on EBL remains controversial, and no guidelines on its use exist.^9, 12-16^

Different techniques for preoperative embolization can be employed, with the goal typically being to achieve complete devascularization of the tumour. When this is achieved, intraoperative bleeding can be significantly reduced compared to controls who do not receive preoperative embolization.^1, 4, 9, 14^ However, a recent meta-analysis^1^ found that EBL has been declining in more contemporary studies without corresponding changes in complete embolization rates, signaling the multifactorial nature of EBL. Some of these confounding factors, such as tumour vascularity and location, require further research. EBL reduction is primarily realized in the intralesional operative setting,^5^ but en bloc surgeries can also benefit from embolization^3^ via devascularization of vessels at the surgical margins, facilitating tumour resection. Although some have seen reduced EBL in en bloc cases as well,^17^ at our centre embolization serves primarily as a technical aid to surgeons.

Given the ambiguity in the literature, this retrospective study seeks to explore the potential effect of embolization completeness on clinical outcomes. We hypothesize that patients undergoing intralesional surgery will have better surgical outcomes including reduced EBL, improved neurological status, and less complications with complete as opposed to partial or near complete embolization. As en bloc surgeries often involve the dissection of large amounts of non-embolized tissue, they are considered a separate cohort of patients and we expect the completeness of embolization in them to have no measurable effect on EBL or other clinical outcomes.

## Methods

The study was approved by the UBC clinical research ethics board (reference # H16-02462). All records of preoperative spine tumour embolization procedures performed by a single operator over 11 years (2007-2018) at Vancouver General Hospital were assessed consecutively.

Patients were referred to the Spine Centre as outpatients, or occasionally as urgent referrals if they had significant clinical morbidity associated with their tumours (e.g., acute/worsening radiculopathy, spinal cord compression, cauda equina syndrome). At our centre, pre-operative embolization is strongly desired for all patients undergoing spine tumour resection, even those considered low risk for intraoperative bleeding, due to operative benefits such as lesion localization and management of segmental arteries. We selected embolization procedures performed by a single neurointerventionist (corresponding author, 16 years of experience) to eliminate interoperator variability.

Embolization was characterized by route of access as well as the type of embolic agent used. Vascularity was assessed by the corresponding author through retroactive visual analysis of the angiographic images obtained during embolization, blinded to outcomes. Tumours were graded as hypovascular or mild, moderate, or highly hypervascular. Devascularization was graded as: complete (100%), near complete (>90%), and partial (>70%) visualized angiographic tumour blush reduction, with the latter two grouped as “incomplete” for statistical purposes. Tumours were characterized by their location in the spine and their vascularity. Location was grouped as upper thoracic (T1-6), thoracolumbar (T7-L5), and sacral. Cervical spine tumours (n=4) were excluded from the analysis due to the unique complexity of these cases.

Clinical outcome data collected included EBL, neurological status change, and presence of any surgical or embolic complications. Estimated blood loss was determined by taking vacuumed blood volumes when autologous transfusion was used (17 operations), or otherwise taking the highest recorded estimate from the operative note or clinical progress notes. Neurological status change was based on neurological physical exams performed by the surgical care team, with the last available preoperative exam compared to the last available postoperative exam.

To minimize the effects of interrater variability, a change was defined as a difference of two or more units on the standard motor scale (from 1-5) or any change in bowel/bladder function. Any explicit statement in the chart noting improvement or worsening of neuromotor function from a health care professional was also considered a change, ensuring that patients’ subjective statements were not used. Finally, surgical and embolization-related complications were noted. These were obtained from the clinical chart and included inadvertent dissection or non-target embolization events, prolonged/copious operative bleeding, inadvertent vascular injury, and intracranial hypotension. Operative duration was obtained from nursing logs, gathered primarily as a control variable to account for the heterogeneity between surgeries.

### Statistical analyses

A multivariable negative binomial regression model was fit to examine the association between EBL and vascularity (categorical: Hypovascular / Mild hypervascularity / Moderate hypervascularity / High hypervascularity), location (categorical: Thoracolumbar / Upper Thoracic / Sacral), completeness of embolization (binary: Yes / No), type (binary: Intralesional / En bloc) and operative duration (continuous). All statistical analyses were performed in R.^18^ Statistical significance was set to *P* < 0.05.

## Results

The review found 40 consecutive patients (aged 21 - 83, mean 58; 9 females, 31 males) with 42 embolization procedures, of which surgery was done en bloc in 22 cases and intralesional in the remaining 20. Two patients had a second procedure; one involved a different spine level, while another had a recurrence after nine years. There was a thoracolumbar predominance in tumour distribution, with particles and coils being the commonest embolic agents used, and complete embolization achieved in the majority of cases (Table 1). A small number of cases were done on an urgent basis (n=9), meaning the patient presented to the emergency department or had rapid neurological decline preceding surgery. There was a diverse mix of benign and malignant spine tumours (data not shown), with the largest group being renal cell carcinoma metastases (RCC, n = 13).

**Table 1:**
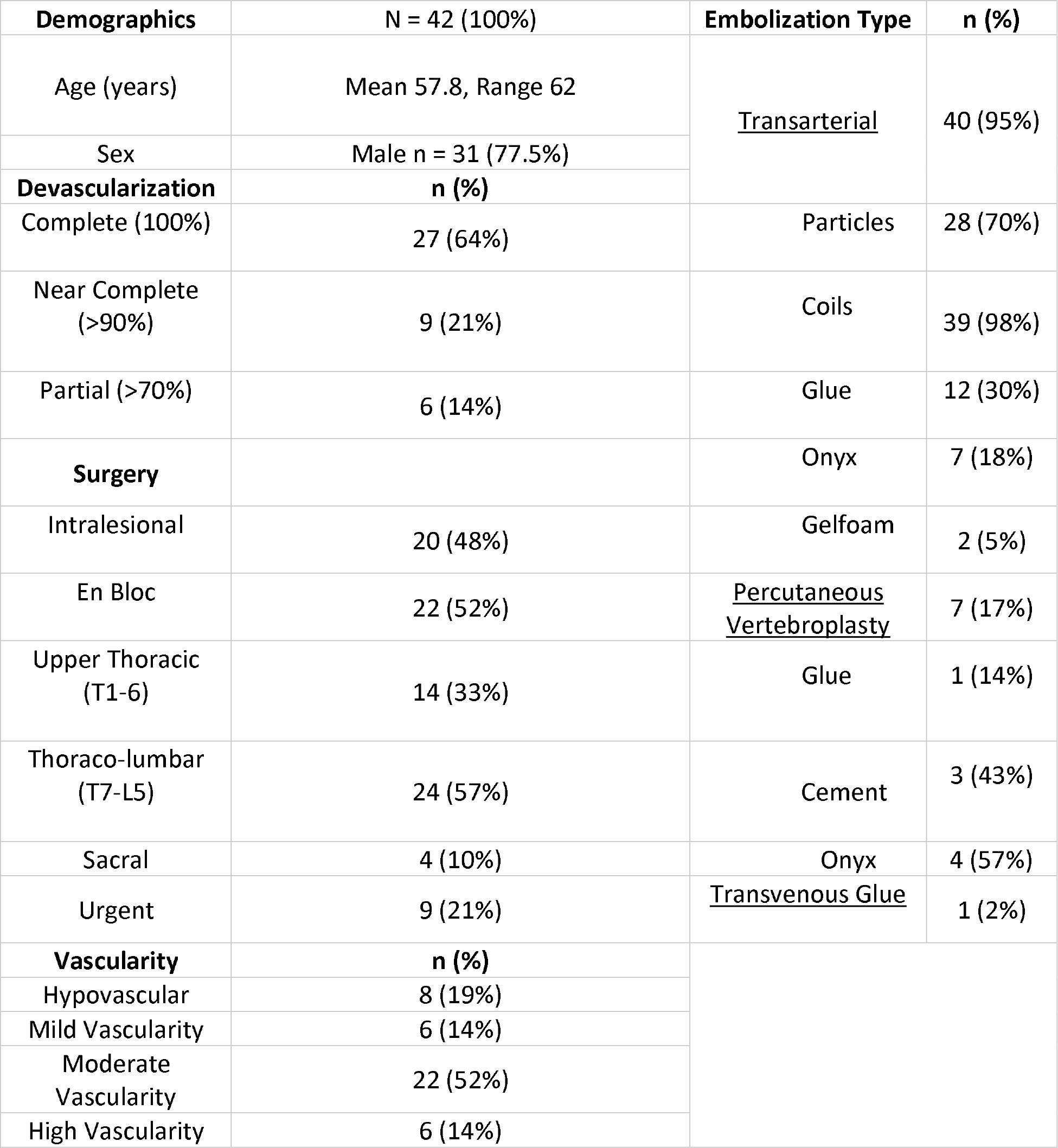
Baseline Data.

Clinical outcome measures are shown in Table 2, with the statistical models outlined in Tables 3 (entire cohort) and 4 (intralesional surgery subgroup).

**Table 2:** Clinical Outcomes.

| (n) | EBL, mean (95% CI) mL | Negative Neurologic Change, n (%) | Complications, n (%) | Negative Neurological Change or Complications, n (%) | Operative Duration, mean (95% CI) minutes |
| --- | --- | --- | --- | --- | --- |
| **En bloc (22)** | 2519*<br>(1782 – 3255) | 6 (27) | 5 (23) | 9 (41) | 705<br>(585 – 826) |
| **Complete (18)** | 2529<br>(1657 – 3401) | 6 (33) | 4 (22) | 8 (44) | 742<br>(600 – 883) |
| **Incomplete (4)** | 2475<br>(288 – 4662) | 0 | 1 (25) | 1 (25) | 543<br>(330 – 756) |
| **Intralesional (20)** | 1133*<br>(660 – 1606) | 1 (5) | 3 (15) | 4 (20) | 310<br>(265 – 354) |
| **Complete (9)** | 772**<br>(360 – 1185) | 0 | 0 | 0 | 317<br>(262 – 371) |
| **Incomplete (11)** | 1428**<br>(606 – 2250) | 1 (9) | 3 (27) | 4 (36) | 304<br>(228 – 380) |
| **High-Vasc (4)** | 1834^<br>(-680 – 4349) | 0 | 1 (25) | 1 (25) | 309<br>(261 – 356) |
| **Mod-Vasc (12)** | 1073<br>(512 – 1633) | 0 | 2 (17) | 2 (17) | 265<br>(216 – 313) |
| **Mild-Vasc (4)** | 613^<br>(143 – 1083) | 1 (25) | 0 | 1 (25) | 445<br>(376 – 515) |
| **T1-6 (11)** | 1085<br>(425 – 1746) | 0 | 3 (27) | 3 (27) | 313<br>(256 – 369) |
| **T7-L5 (8)** | 1278<br>(322 – 2233) | 1 (13) | 0 | 1 (13) | 312<br>(214 – 411) |
| **Sacral (1)** | 500 | 0 | 0 | 0 | 255 |
| **Entire Cohort (42)** | 1859<br>(1378 – 2339) | 7 (17) | 8 (19) | 13 (31) | 517<br>(428 – 606) |
| **High-Vasc (6)** | 2852^^**~<br>(718 – 4986) | 1 (17) | 2 (33) | 2 (33) | 375<br>(218 – 532) |
| **Mod-Vasc (22)** | 1585~<br>(1062 – 2108) | 2 (9) | 3 (14) | 4 (18) | 443<br>(342 – 545) |
| **Mild-Vasc (6)** | 1480**<br>(-571 – 3531) | 1 (17) | 1 (17) | 2 (33) | 531<br>(271 – 790) |
| **Hypo-Vasc (8)** | 2151^^<br>(760 – 3542) | 3 (38) | 2 (25) | 5 (63) | 815<br>(517 – 1114) |
| **T1-6 (14)** | 1257<br>(716 – 1797) | 1 (7) | 3 (21) | 4 (29) | 386<br>(289 – 484) |
| **T7-L5 (24)** | 2116<br>(1411 – 2820) | 5 (21) | 5 (21) | 8 (33) | 573<br>(452 – 694) |
| **Sacral (4)** | 2428<br>(-1021 – 5877) | 1 (25) | 0 | 1 (25) | 638<br>(-147 – 1424) |

\* P = 0.049 \*\* P < 0.001 ^ P = 0.005 ^^ P=0.004 ~ P = 0.022

**Table 3:** Multivariable negative binomial regression model examining the association between blood loss and vascularity, location, completeness of embolization, surgical type and duration in the entire cohort (n = 42).

|  | ESTIMATE | STANDARD ERROR | P-VALUE |
| --- | --- | --- | --- |
| **INTERCEPT** | 7.46 | 0.41 | <0.001 |
| **VASCULARITY** |  |  |  |
| HYPOVASCULAR | -1.05 | 0.36 | 0.004 |
| MILD HYPERVASCULARITY | -1.12 | 0.33 | <0.001 |
| MODERATE HYPERVASCULARITY | -0.61 | 0.27 | 0.022 |
| HIGH HYPERVASCULARITY | REF | REF | REF |
| **LOCATION** |  |  |  |
| UPPER THORACIC | -0.04 | 0.21 | 0.857 |
| THORACOLUMBAR | REF | REF | REF |
| SACRAL | 0.13 | 0.31 | 0.669 |
| **EMBOLIZATION: INCOMPLETE** | 0.37 | 0.21 | 0.084 |
| **SURGERY TYPE: INTRALESIONAL** | -0.52 | 0.26 | 0.049 |
| **OPERATIVE DURATION** | 0.0014 | 0.00046 | 0.003 |

**Table 4:** Multivariable negative binomial regression model examining the association between blood loss and vascularity, location, completeness of embolization, and operative duration in the intralesional surgery group (n=20; 48%).

|  | ESTIMATE | STANDARD ERROR | P-VALUE |
| --- | --- | --- | --- |
| **INTERCEPT** | 5.93 | 0.82 | <0.001 |
| **VASCULARITY** |  |  |  |
| MILD HYPERVASCULARITY | -1.33 | 0.47 | 0.005 |
| MODERATE HYPERVASCULARITY | 0.12 | 0.35 | 0.722 |
| HIGH HYPERVASCULARITY | REF | REF | REF |
| **LOCATION** |  |  |  |
| UPPER THORACIC | -0.26 | 0.24 | 0.277 |
| THORACOLUMBAR | REF | REF | REF |
| SACRAL | -1.53 | 0.58 | 0.009 |
| **EMBOLIZATION: INCOMPLETE** | 1.02 | 0.28 | <0.001 |
| **OPERATIVE DURATION** | 0.0026 | 0.0021 | 0.221 |

### Entire Cohort

EBL was reduced in hypovascular, mild, and moderately hypervascular tumours when compared to tumours with high hypervascularity (P=0.004, P<0.001, and P=0.022 respectively). Intralesional surgery was associated with reduced EBL relative to en bloc surgery (p=0.049). An increase in operative duration was associated with an increased EBL (p=0.003). Location of tumours and completeness of embolization were not associated with EBL in the entire cohort sample.

### Intralesional Surgeries

EBL was reduced in mildly hypervascular tumours when compared to tumours with high hypervascularity (P=0.005). No hypovascular tumours were available in this cohort for comparison. In addition, the single sacral tumour in this cohort was associated with reduced EBL when compared to lesions in the thoracolumbar region (p=0.009). Incomplete embolization was associated with increased EBL relative to complete embolization (p<0.001). Operative duration was not associated with EBL in the intralesional surgery sample.

There were no statistically significant differences in neurological outcomes or complications between the complete and incomplete embolization groups within either cohort (Table 2).

## Discussion

Our main finding was that the completeness of tumour embolization correlates with blood loss in intralesional surgeries, providing preliminary evidence in support of our hypothesis. This has been a conclusion of several similar studies.^10, 11^ While other authors have not found a benefit to greater devascularization,^12-16^ limited small sample sizes and confounding factors such as surgical technique may have concealed a potential effect. Furthermore, the heterogeneity between studies limits direct comparisons, as for example the definition of complete devascularization varies.

This study also re-affirms the blood-loss reduction benefit of pre-operative spine tumour embolization in general. Our overall average EBL of around two litres is similar to those in other studies, with overlapping 95% confidence intervals.^2-5, 7^ The increased blood loss seen with en bloc cases is also comparable to the literature^17^ and can be explained by the nature of these surgeries, which includes dissection and ligation of tissues that cannot be embolized, larger surgical exposures and more complex hardware reconstructions necessitating longer operative durations. In intralesional surgeries, two older studies with controls showed an average EBL of 5 and 6.7 L without versus 1.5 and 4.3 L with embolization respectively,^4, 5^ demonstrating the benefit of this procedure. Our intralesional group EBL averaged 1.1 L, concordant with earlier studies, and any further reduction in EBL achieved in our study is likely secondary to advances in embolization and surgical techniques.

We controlled for operative duration, which correlated with EBL in the entire cohort, to capture much of the heterogeneity between cases including tumour size, degree of canal compromise, type of stabilization and extent of exposure. Interestingly, surgical duration was comparable in both complete and incomplete embolization groups, suggesting that any potential benefit this procedure has in expediting surgery may not depend on extent of embolization, or may be veiled by other factors.

Surprisingly, the tumour spinal level appears unrelated to EBL in our study. This despite upper thoracic lesions being more difficult to embolize and associated with more technical operations. The finding of lower EBL in the intralesional sacral group (n=1) is unlikely to be representative. Additionally, highly vascular tumours correlated with higher EBL despite embolization, which may relate to the effect of incomplete devascularization or peripheral angiogenesis surrounding the tumour.

It is important to note that EBL is a very crude measure and does not necessarily best capture the benefit of embolization as there are many factors which can account for blood loss. The consensus from surgeons at our centre is that ‘complete’ embolization is very helpful in facilitating surgery, particularly in intralesional cases where tumor devascularization is directly observed. With respect to en bloc procedures, as these are almost always large operations, substantial EBL is expected. However, again, embolization is felt to facilitate the steps in performing the surgery in a binary manner with no importance attributed to the degree of embolization achieved, also suggested by our data.

In terms of other clinical outcomes, although numerically there was a higher incidence of neurological decline or complications in intralesional surgeries between incomplete (36%) versus complete (0%) embolization, our study did not show a statistical difference. The reduced EBL potentially conferred by complete embolization would be expected to positively impact neurological outcomes. However, as the incidence of negative outcomes is relatively sparce, larger sample sizes are needed to show any potential differences. Comparison of our neurological outcomes and complications with those of the literature is difficult given the heterogeneity in the patients with respect to their tumours, pre-surgical medical status, complexity of surgery, post-surgical recovery, and method of neurological assessment used.

Finally, given the observed potential benefit of achieving complete embolization, techniques that maximize full devascularization of tumours are suggested for intralesional surgeries if they can be safely deployed. At our centre, progression towards increasing use of percutaneous embolization has improved our ability to safely increase complete embolization rates. A full discussion of this is previously described.^19^

## Limitations

Our retrospective chart review has several limitations. First, we had to rely on inconsistencies in clinical documentation, including measurement of EBL as well as the patient’s neurological status. We tried to minimize this heterogeneity by using autologous blood transfusion data as much as possible to standardize EBL and by cross-referencing several neurological exams at each time point. Second, our sample size of 42 embolization cases limited the statistical power of our analysis. A slight bias may have been introduced by treating second operations as independent cases, although this was limited to only two patients. Third, our population was predominantly male (78%), which limits the generalizability of our findings as it has been shown that males have poorer neurological outcomes compared to females.^6^

## Conclusion

This study provides early evidence for our hypothesis given that complete as opposed to incomplete embolization of tumours correlated with reduced blood loss in intralesional surgeries. This observation has also been endorsed by the spine surgeons at our centre. However, randomized control trials are needed with larger cohorts to confirm our hypothesis and better ascertain other potential clinical benefits of pre-operative tumour embolization beyond EBL.

## Data Availability

All data produced in the present study are available upon reasonable request to the authors

## Abbreviations

EBL: estimated blood loss

